# Machine learning-based prediction of future dementia using routine clinical MRI brain scans and healthcare data

**DOI:** 10.1101/2025.11.12.25340070

**Authors:** Parminder Singh Reel, Salim Al-Wasity, Craig Edwards, Smarti Reel, Esma Mansouri-Benssassi, Szabolcs Suveges, Muthu Rama Krishnan Mookiah, Susan Krueger, Emanuele Trucco, Emily Jefferson, Alexander Doney, J. Douglas Steele

## Abstract

**Importance:** Early identification of dementia risk is essential for preventive care and timely enrolment into disease-modifying interventions. Current approaches rely on costly, invasive, or research-only methods not feasible at scale within public health systems.

**Objective:** To test whether routinely acquired NHS brain MRI scans can be used to predict future dementia diagnosis and whether confidence-based stratification improves prediction reliability and clinical interpretability.

**Design, Setting, and Participants:** Retrospective case-control study conducted entirely within a secure NHS Trusted Research Environment. Routine T1-weighted MRI brain scans were linked with electronic health records for participants from Tayside and Fife, Scotland. The study included 259 individuals who subsequently developed dementia and 259 age- and sex-matched controls. Data were processed and modelled between January and June 2025.

**Exposure:** MRI-derived structural brain features analysed using a support-vector-machine model with nested cross-validation and distance-from-hyperplane (DFH) confidence calibration.

**Main Outcomes and Measures:** Primary outcomes were prediction accuracy and area under the receiver-operating-characteristic curve (AUC). Secondary analyses assessed DFH-stratified performance and time from scan to clinical diagnosis.

**Results:** Confidence-based filtering identified a high-confidence subgroup (≈35% of scans) with ≈80% accuracy. Overall, the model predicted future dementia up to five years before first recorded NHS diagnosis, achieving 66.8% accuracy (AUC = 0.71). Model sensitivity increased for shorter time-to-diagnosis intervals. Analyses were generalisable across heterogeneous routine NHS scanners and datasets.

**Conclusions and Relevance:** This study provides, to our knowledge, the first demonstration that routinely collected NHS MRI data can predict future dementia years before clinical diagnosis. Incorporating confidence calibration transforms a standard classifier into a safety-aware, clinically interpretable framework, supporting scalable early detection, risk stratification, and recruitment to preventive or disease-modifying trials across population health systems.

**Key Points:** *Question:* Can routinely acquired NHS magnetic resonance imaging (MRI) brain scans and linked health records be used to predict future dementia, with confidence calibration suitable for clinical and population-level use?

*Findings:* In this retrospective case-control study of 518 individuals, a support-vector-machine model trained on routine NHS MRI data predicted future dementia up to five years before diagnosis. Confidence-based stratification identified a high-confidence subgroup (∼35% of scans) with ∼80% accuracy.

*Meaning:* Routine NHS imaging data can be transformed into a population-scale, privacy-preserving early detection framework, where confidence calibration allows safe, clinically interpretable application for preventive care and research recruitment.

## Introduction

Dementia poses one of the most pressing global health challenges, affecting more than 55 million people worldwide and projected to nearly triple by 2050 as populations age^1,2^. In the UK, dementia care already exceeds £34 billion annually^3^, imposing major economic, healthcare, and social burdens. Beyond these costs, the disease carries profound personal and societal consequences^4^. Alzheimer’s disease (AD) accounts for 60-80% of cases^5^, followed by vascular dementia (∼20%), and despite decades of research, effective disease-modifying treatments remain limited.

Once clinical symptoms emerge, substantial and irreversible neuronal loss has typically occurred, rendering interventions less effective. Recent advances in disease-modifying therapies for AD, such as amyloid-targeting monoclonal antibodies, highlight the importance of identifying individuals at pre-symptomatic or prodromal stages^6,7^. However, early diagnosis remains difficult: current methods rely on specialized cognitive assessments and invasive or costly biomarkers, including PET imaging and cerebrospinal fluid analyses^8^, which limit scalability for routine use. Although blood-based biomarkers show promise^9^, they have yet to achieve widespread clinical implementation.

Magnetic resonance imaging (MRI) provides a rich, non-invasive source of information about brain structure and health. Machine learning (ML) applied to MRI has shown considerable promise for detecting subtle neurodegenerative patterns that precede dementia symptoms^10–12^. Deep learning and support-vector-machine (SVM) models can distinguish Alzheimer’s disease, mild cognitive impairment, and healthy aging with high accuracy^13^. However, most models rely on research-grade imaging datasets that are carefully curated and homogeneous. These datasets do not represent real-world healthcare populations, which limits the generalizability of such models and their potential for deployment within health systems.

Dementia therefore represents both an urgent clinical need and an opportunity for scalable, data-driven risk prediction. Routinely acquired National Health Service (NHS) brain scans, if effectively leveraged, could enable population-wide dementia risk modelling within privacy-preserving Trusted Research Environments (TREs). Unlike research datasets, these scans capture the heterogeneity of clinical imaging and could support large-scale, real-world implementation.

The present study addresses this opportunity by demonstrating, for the first time, that future dementia can be predicted up to five years before its first recorded NHS diagnosis using routinely collected MRI data. We further introduce a distance-from-hyperplane (DFH) confidence calibration method that quantifies model certainty. This approach transforms a conventional classifier into a clinically interpretable and safety-aware decision-support tool suitable for early intervention triage and precision trial recruitment.

Leveraging longitudinal healthcare data from the NHS provides a transformative approach to dementia research and patient care^14^, enabling long-term tracking of disease risk and progression^15^. Integrating such data with routine imaging, for example through national repositories such as the Scottish Medical Imaging (SMI) resource^14^, creates a strong foundation for developing predictive models that reflect real-world practice. However, this approach introduces challenges that include diagnostic labelling, data harmonization, quality control, and secure computation. These requirements must be met within Trusted Research Environments, or “Safe Havens,” that safeguard patient privacy^16,17^.

To address these challenges, we conducted a retrospective, case-control study of routinely acquired NHS brain MRI scans linked to longitudinal electronic health records within a secure TRE. Using a nested cross-validated SVM framework, we trained and tested models to distinguish individuals who later developed dementia from matched controls with no dementia diagnosis. Structural brain variability was captured through established neuroanatomical deformation metrics derived from standard clinical T1-weighted images, allowing analysis across heterogeneous scanners typical of NHS practice.

To enhance clinical reliability, DFH-based stratification was applied to quantify model confidence and enable selective prediction of high-certainty cases. This strategy provides an interpretable balance between predictive precision and population coverage. The findings demonstrate the feasibility of developing population-scalable dementia risk prediction tools using routine healthcare imaging data.

## Methods

### Data Sources

We combined two previously established research healthcare datasets managed by Health Informatics Centre (HIC) at the University of Dundee which have been described previously: GoDARTS (Genetics of Diabetes Audit and Research in Tayside Scotland)^18^ and SHARE (Scottish Health Research register)^19^. All available MRI brain scans collected as part of routine NHS medical practice for Tayside and Fife participants in these studies were obtained from the SMI^14^. The use of data by this project was covered by HICs ethical approval 18/ES/0126, in accordance with conditions set by the East of Scotland Research Ethics Committee, and the approval of the NHS Caldicott Guardians. All SHARE participants consent to the access and use of their electronic health records for eligible research projects such as this one, scrutinised and approved by the SHARE access committee.

### Workflow Design

Figure 1 illustrates an end-to-end six-staged workflow: accessing routinely collected healthcare data and images, constructing study populations, image data curation, pre-processing and support vector machine (SVM)-based ML pipeline and result egress within the TRE environment. Each of these stages is described below.

**Figure 1:**
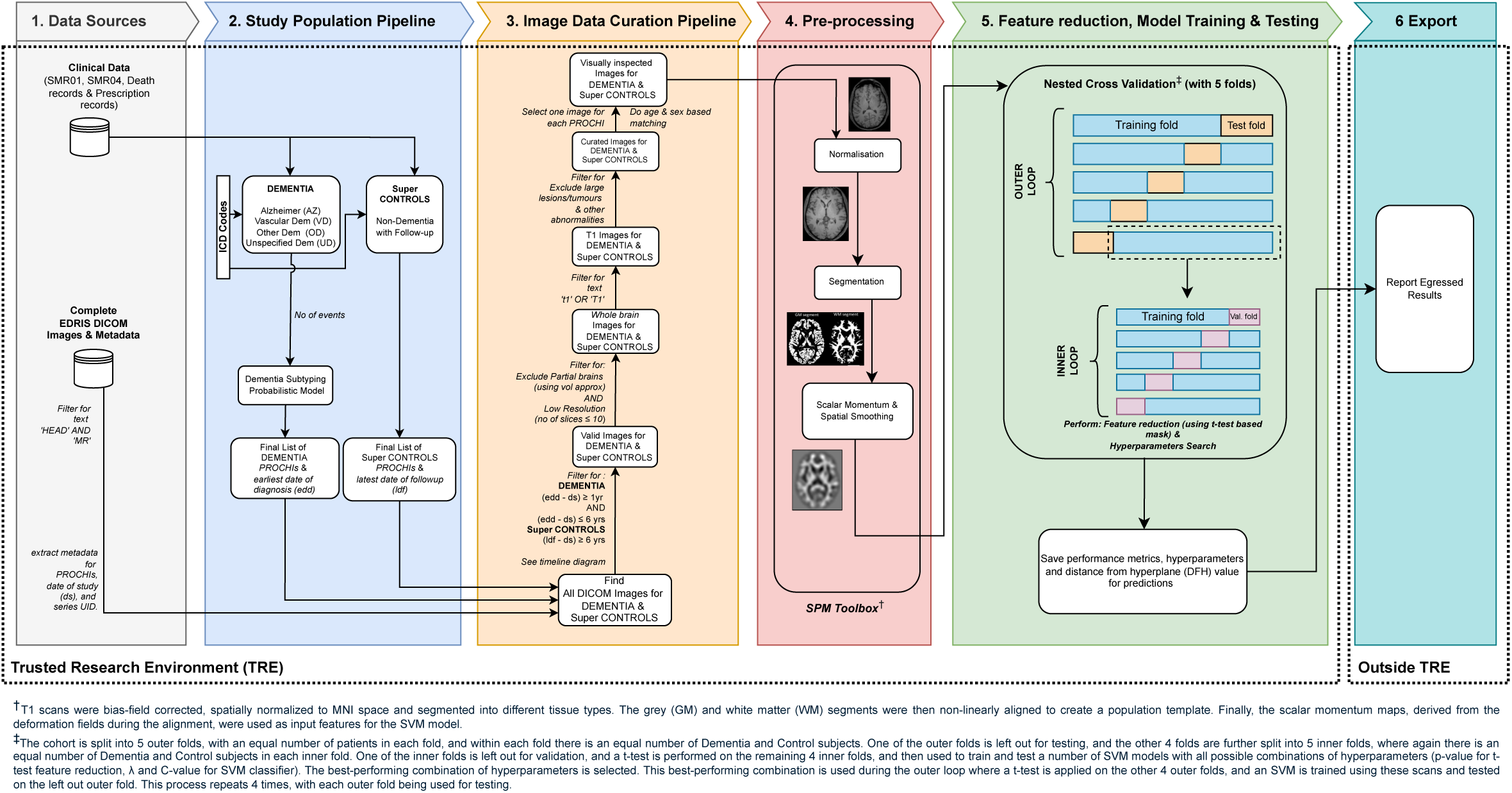
End-to-end six-staged workflow for using routine clinical MRI brain scans and linked prospective electronic medical records (EMR) to predict future dementia using SVM Approach.

### Study Population Pipeline

Using our validated electronic medical record algorithm^20^ cases of Alzheimer’s dementia (AD) and vascular dementia (VD) cases were identified. In addition, two other groups of cases were identified: “Other dementias” (OD) that include a variety of less common types of dementia, e.g., Lewy body dementia and frontotemporal dementia and “Unspecified dementia” (UD), when the algorithm could not definitively classify the dementia type. Non-dementia controls were identified as patients who did not ever develop dementia. For further details of dementia cases and non-dementia controls definitions see eMethod 1 and eTable 1.

### Image Data Curation Pipeline (IDCP)

This pipeline incorporates a sequential series of filtering steps applied to the image files and DICOM meta-data linked to the study population. Its purpose was to select MRI scans suitable for inclusion in the ML training. Metadata were first filtered for ‘T1’, ‘MR’, and ‘Head’ tags and time window applied: 1-6 years pre-diagnosis for cases, and a minimum of 6 years follow-up after the date of scan or non-dementia death for controls (see eFigure 1). A custom annotation tool (see eFigure 2) enabled manual review in orthogonal planes to exclude scans with artefacts (e.g., partial brains using volume calculator, see eFigure 3) or anatomical lesions). Age and sex were matched across groups. Final scans were converted to NIfTI format. For further details of IDCP see eMethod 2.

### Preprocessing

All T1-weighted magnetic resonance scans from IDCP were pre-processed using SPM12^21^, running under MATLAB^TM^ R2021a^22^. Scans underwent bias-field correction, spatial normalization to the MNI template, and tissue segmentation into grey matter (GM), white matter (WM), and cerebrospinal fluid (CSF) as shown in Figure 1. Nonlinear within-subject registration was then performed using the SPM12 Geodesic Shooting Toolbox^23^, iteratively refining a population template from segmented GM and WM images. Deformations were computed and applied until a stable, representative template was achieved. Finally, ‘scalar momentum’^24^ was calculated to capture deformation fields, tissue density differences, and morphological variations that provides a rich structural representation to enhance machine learning performance. For further details see eMethod 3.

### Nested Cross-validation

Nested cross-validation was used with the dataset split into five outer folds, with one-fold for testing and the remaining four for ML model training. Within the training set, five inner folds enabled hyperparameter tuning. Feature reduction was performed using a variable-threshold t-test approach within a multivariate pattern analysis framework as discussed below:

#### Feature Reduction

To prevent overfitting from high-dimensional data, this study used voxel-wise feature reduction via a two-sample t-test within a nested cross-validation framework. This method selects informative voxels while discarding noisy or redundant voxels, enhancing classification performance and reducing computational cost. The t-test was applied only to training data to avoid data leakage, produced a t-map indicating voxel significance between groups, avoiding double dipping. A binary t-mask was created using an optimal p-value threshold, selected at the same time as optimal SVM hyperparameters. This t-mask was applied to the test set to assess model performance across thresholds, improving generalisability. An additional benefit of using this method was that it explicitly identifies brain regions which were used for predictions, thereby enhancing model transparency. For further details see eMethod 4.

#### Multi-Variate Pattern Analysis

Radial Basis Function (RBF) SVM as implemented in MATLAB^22^ was chosen for its effectiveness with high-dimensional neuroimaging data and superior performance on small datasets compared to deep learning models^13^. The nested cross-validation framework optimized key hyperparameters: box-constraint “C”, and kernel width “λ”, at the same time as p-value threshold for t-test threshold selection, using predefined ranges and a cloud-based SLURM computing (see eMethod 5). Feature reduction was applied only to training data, generating t-masks for voxel selection, which were then fixed for testing. During training model performance was evaluated on the inner validation folds, and the best hyperparameter set was used for the outer fold as shown in Figure 1. Final accuracy was averaged across all outer folds.

#### Improving Classification Reliability with SVM DFH (exploratory analysis)

In this exploratory analysis, an SVM with a “reject” option was used to enhance classification reliability by excluding “low-confidence” predictions, defined as Distance From the Hyperplane (DFH)^25^. This is especially important in medical settings, where misclassification can have serious consequences^26^. DFH provides a measure of prediction confidence, helping clinicians assess individual patient risk of future dementia. Accuracy was calculated by rejecting predictions with DFH below a set threshold, focusing on more confidently classified cases^27,28^. Performance metrics were evaluated across DFH thresholds from 0 to 1 in 0.05 increments, allowing analysis of trade-offs between confidence, accuracy, and scan coverage.

### Export

All results were securely stored within the TRE and submitted for egress. The TRE administrator conducted disclosure checks^29^, redacting sensitive data. Approved result files were then used for reporting in this study.

## Results

### Cohort Identification Pipeline

Overall, 3,477 patients (AZ [*n*=1,858], VD [*n*=800], UD [*n*=617], and OD [*n*=202]) who had a diagnosis of dementia in their medical records were identified within the source population (see Figure 2 and eMethod 6), and 99,105 patients who never developed such a diagnosis.

**Figure 2:**
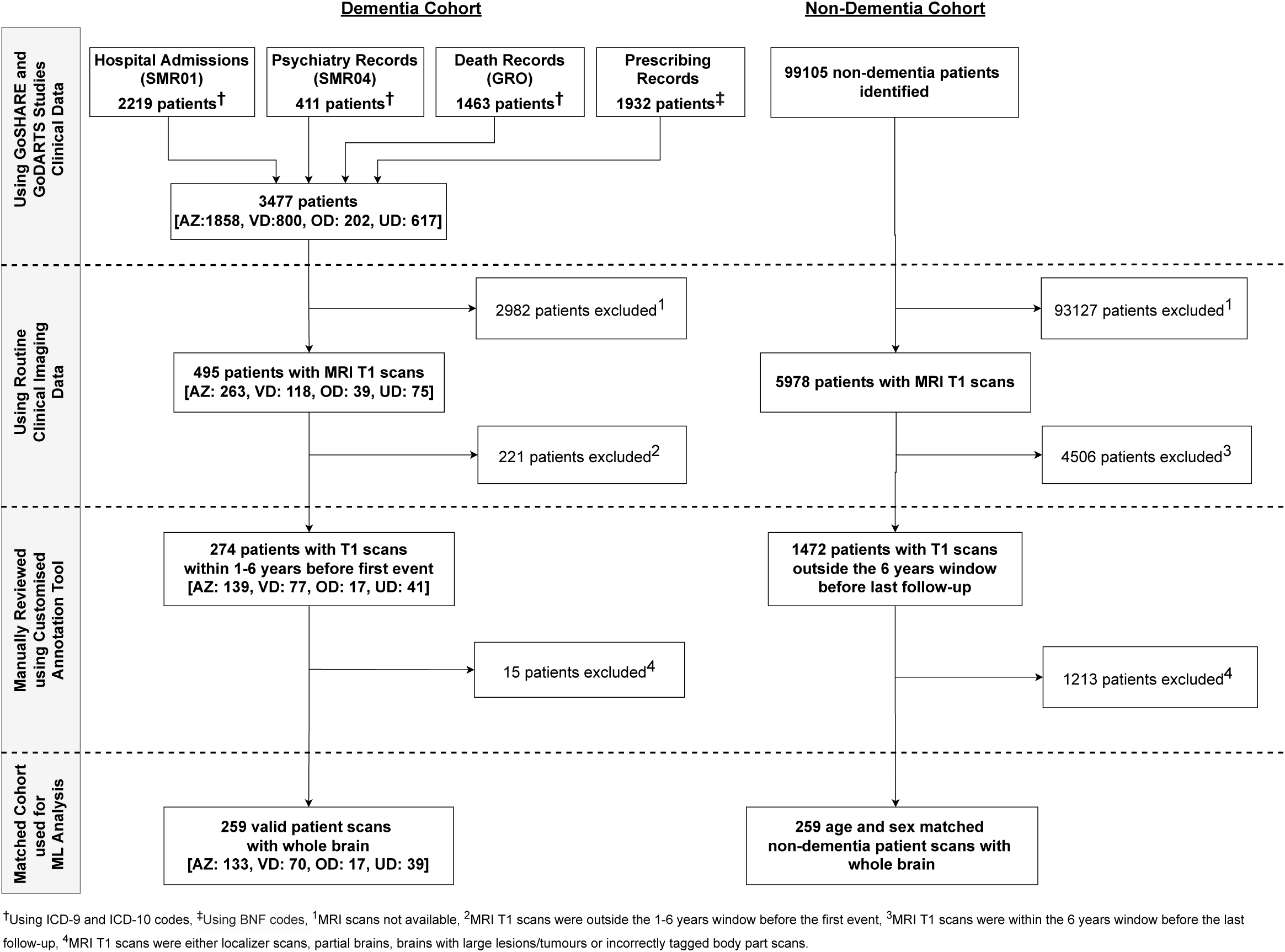
Patient cohort identification and data linkage utilising available EHR and imaging data.

### Study population and Image Data Curation Pipeline

In total, 495 dementia cases (AZ [*n*=263], VD [*n*=118], UD [*n*=75], and OD [*n*=39]) had linked T1 MRI brain scans in routine clinical imaging data. Finally, 259 valid case scans (AZ [*n*=133], VD [*n*=70], UD [*n*=39], and OD [*n*=17]) were identified after IDCP evaluation. These were age and sex matched with 259 control scans (manually reviewed and validated for any exclusions post-age and sex matching), corresponding to valid case scans. As expected, NHS scans were unlike research scans as they had been acquired using heterogenous scanner parameters (see details in eFigure 4). Table 1 provides the population characteristics for this matched study cohort.

**Table 1:**
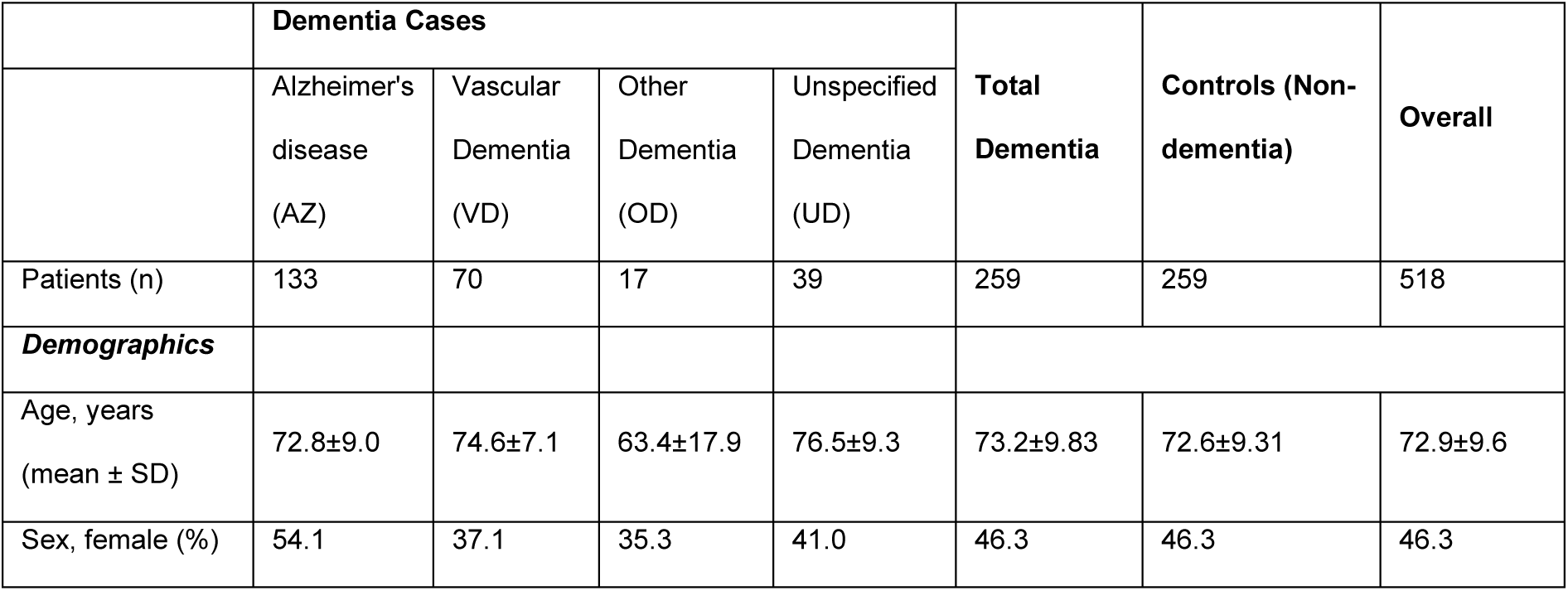
Demographic and clinical characteristics of the study groups.

### Feature Reduction

A higher number of most significant voxels were selected as the p-value thresholds (*p_Thres*) gradually increased during the feature reduction in 5-fold nested cross validation as illustrated in eFigure 5. The final selected t-mask regions used for the outer folds were very similar. Figure 3 (A) shows the final selected t-mask used for each outer fold overlaid on publicly accessible brain scan conforming to standard Montreal Neurological Institute (MNI) anatomical space.

**Figure 3:**
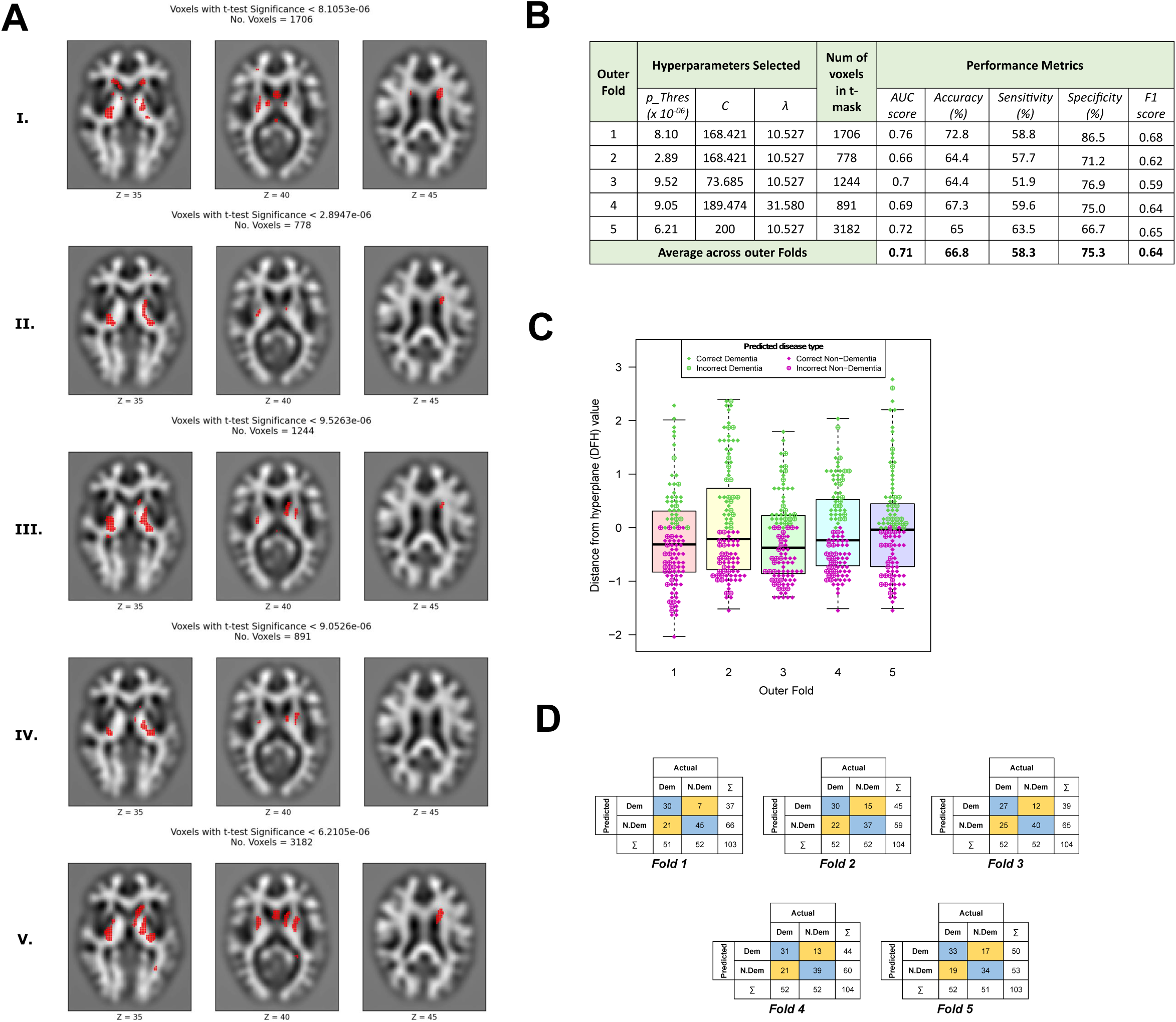
(A) Significant Voxels Overlaid on COLIN Brain Scan for (I) Fold 1, (II) Fold 2, (III) Fold 3, (IV) Fold 4 and (V) Fold 5. The COLIN Brain Scan was pre-processed by segmenting into grey matter and smoothed using scalar momentum. The three scans in each row display different axial slices, inferior to superior across from left to right. (B) Overall summary of performance metrics; (C) Distribution of DTH value for prediction outcomes and (D) Confusion matrices across 5-fold nested cross-validation

### MVPA Performance

#### Analysis 1: All Data

The RBF-SVM model performance across five outer folds of the nested cross-validation demonstrated an average AUC score of 0.71 and accuracy of 66.8% in discriminating between scans labelled as future dementia cases and controls as illustrated in Figure 3 (B). Sensitivity, specificity and F1 score averaged 58.3%, 75.3% and 0.64 respectively, indicating a slightly better ability to correctly identify true positives than true negatives. The number of voxels selected by the t-mask varied widely across folds (ranging from 778 to 3182), reflecting variability in the feature reduction process. Selected hyperparameters also varied, with *p_Thres* ranging from *2.89 to 9.52 (×10⁻⁶)*, C values from *73.68* to *200*, and λ values from *10.52* to *31.57*. The full grid search performance across hyperparameters highlight a smooth function as shown in eFigure 6. Overall, these results suggest that while the model maintained consistently moderate performance, there was some variability in optimal parameter settings and feature reduction across folds, as expected. Figure 3 (C) shows the distribution of DFH values for prediction outcomes across each fold.

The confusion matrices across five folds show reasonable consistency in the model’s classification of two classes, with varying strengths across folds as shown in Figure 3 (D). Fold 1 delivers the best balance between sensitivity and specificity, while Folds 2 and 3 show reduced specificity and sensitivity due to increased false predictions. Fold 4 improves this balance slightly, and Fold 5 maintains reasonable accuracy but with a higher false positive rate. Overall, the model performs reliably but exhibits some variability across folds, indicating potential benefits from further tuning or regularization to enhance generalisation.

#### Analysis 2: Filtered Data

Figure 4 (A) shows varying the DFH threshold (ranging from 0 to 1 in 0.05 increments) impacts classification performance, showing a trade-off between sample coverage and classification metrics. As the threshold increases from 0 to 1, the number of classified samples dropped steadily (from 518 to 5), while accuracy and sensitivity generally improved. Early thresholds (0-0.3) prioritise broader classification, with more balanced sensitivity and specificity. From thresholds 0.4 to 0.65, the model achieves a notable balanced sensitivity, specificity while maintaining moderate classification coverage. Beyond 0.7, sensitivity reached 100% as the model became highly conservative, only predicting when very “confident” but at the cost of reduced specificity and sample coverage. At threshold = 1, all classified predictions are correct (100% accuracy), but the sample size was minimal, making performance estimates less reliable. This trade-off illustrates the importance of selecting a threshold that balances confidence with meaningful data coverage for the intended application.

**Figure 4:**
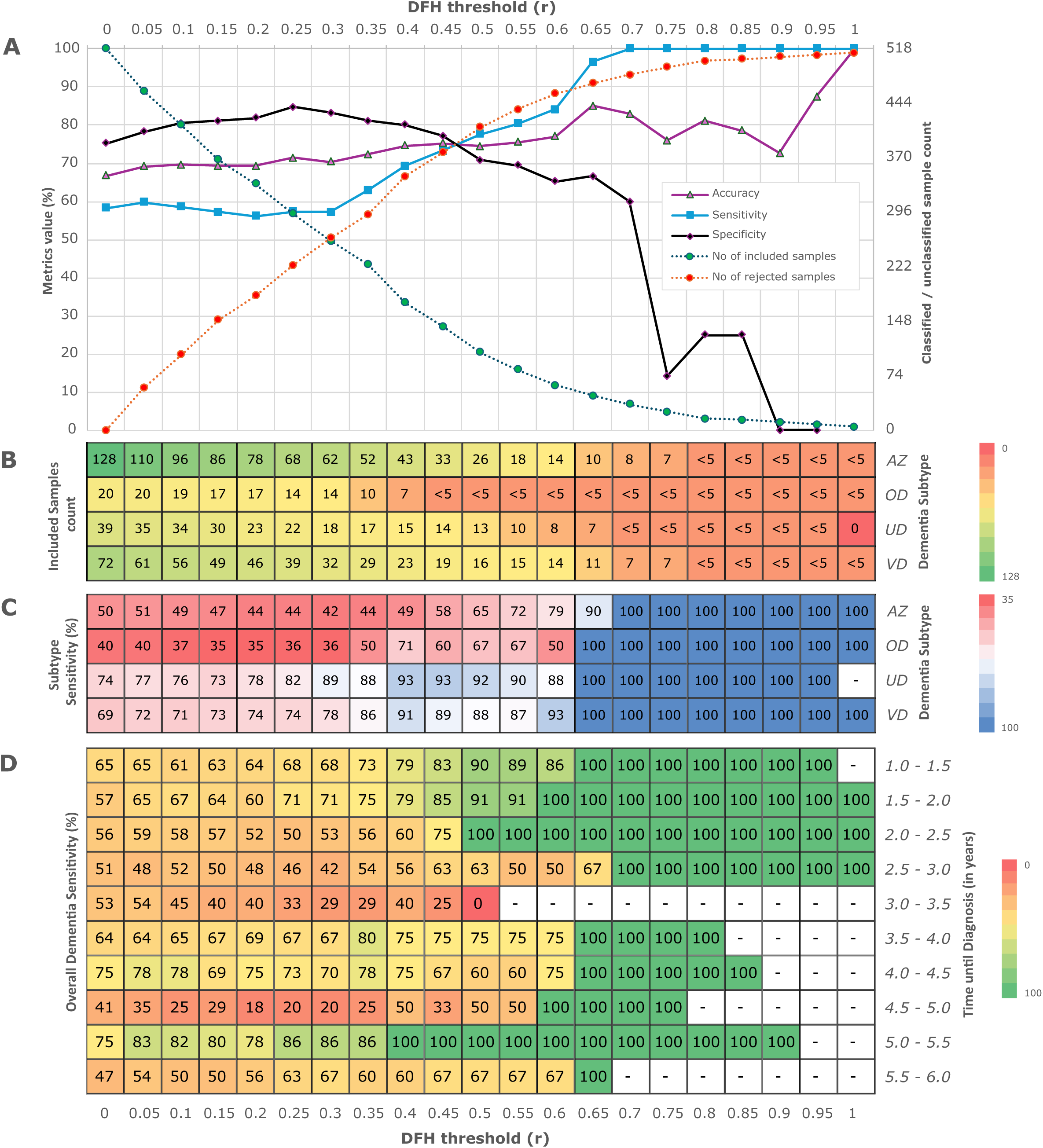
(A) Classification metrics (accuracy, sensitivity, and specificity) with varying DFH threshold, (B) Classified sample count across dementia subtype, (C) Sensitivity (%) across dementia subtypes and (D) overall dementia sensitivity w.r.t time until diagnosis (in years).

#### Prediction of dementia subtypes

The model demonstrated promising results in identifying patients who later developed specific dementia subtypes. Figure 4 (B - C) presents the dementia subtype (AZ, OD, UD, and VD) count and their sensitivity values w.r.t increasing DFH thresholds. As the threshold increases from 0 to 1, the total number of included samples decreases across all subgroups. Correspondingly, sensitivity increases in all categories, indicating improved classification performance with higher-quality inputs. At threshold 0, sensitivities are relatively low to moderate (AZ: 50%, OD: 40%, UD: 74.4%, VD: 69.4%). However, by threshold 0.65, all sensitivity values reach or exceed 90%, with most achieving 100% by 0.7. This trend illustrates a clear trade-off between sample size and model sensitivity: excluding lower-confidence cases enhances performance metrics but reduces the sample size significantly. For thresholds above 0.75, sensitivities remain at 100% for all applicable categories, although the number of retained cases becomes small.

#### Prediction of Time for Individual Patient’s to Develop Dementia

Figure 4 (D) illustrates dementia prediction sensitivity as a function of the time until patients received their diagnosis. Overall, the results demonstrate that applying higher DFH thresholds improves sensitivity across nearly all time intervals, particularly in the earlier stages of disease progression. In the short-term intervals (1.0-2.5 years), sensitivity is already relatively high at baseline (e.g., 1.0-1.5 years: 65%, 1.5-2.0 years: 57%, 2.0-2.5 years: 56%) and increases steadily with higher thresholds, reaching 100% sensitivity by thresholds 0.65-0.75 in most cases. This indicates the model’s strong ability to detect imminent diagnoses under stricter threshold settings. Mid-range intervals (2.5-4.5 years) exhibit greater variability in sensitivity. For example, in the 2.5-3.0 year group, sensitivity fluctuates between 42-63% across thresholds, only reaching 100% at threshold 0.65. The 3.0-3.5 year group shows a marked decline in sensitivity beyond threshold 0.3, with values dropping to 0% at threshold 0.5 and absent thereafter, reflecting reduced model reliability in this interval. In contrast, the 4.0-4.5 year interval maintains relatively high sensitivity (60-78%) at lower thresholds and reaches full sensitivity by 0.6. For longer-term intervals (4.5-6.0 years), sensitivity varies widely. In the 4.5-5.0 year range, values are low at lower thresholds (as low as 18%) but improve to 100% by threshold 0.6. Similarly, the 5.0-5.5 and 5.5-6.0 year groups achieve full sensitivity at thresholds 0.45 and 0.65, respectively, although data becomes sparse or missing beyond those points. Overall, increasing the DFH threshold consistently enhances sensitivity, particularly in early and mid-term diagnosis windows, but often at the expense of sample coverage in longer-term cases.

## Discussion

This study demonstrates that future dementia can be predicted up to five years before the first recorded NHS diagnosis using routinely acquired clinical MRI brain scans linked to electronic health records within a privacy preserving Trusted Research Environment. By analysing data collected during standard clinical care rather than research protocols, we show the feasibility of developing population scalable predictive models capable of operating within real world imaging heterogeneity. This establishes a critical proof of principle that routinely available NHS imaging data can support large scale, prospective dementia risk stratification without the need for new data collection or bespoke imaging sequences. A second major contribution is the introduction of a distance from hyperplane confidence calibration approach, which quantifies the certainty of each individual patient machine learning prediction. This strategy allows the model to prioritise high confidence classifications, achieving approximately 80 percent accuracy for one third of scans while maintaining a consistent baseline performance with AUC equal to 0.71 across the entire dataset. Rather than a post hoc adjustment, distance from hyperplane stratification transforms the classifier into a selective prediction system analogous to clinical reasoning, where uncertain cases can be deferred or referred for further evaluation. This capability enhances interpretability, patient safety, and clinical trust, properties increasingly recognised as essential for the responsible deployment of artificial intelligence systems in healthcare. Taken together, these findings establish three interlocking advances. First, demonstration that routine NHS radiology can underpin national scale dementia risk modelling. Second, evidence that future dementia can be predicted years in advance of NHS diagnosis. Third, introduction of confidence calibrated prediction enabling safe translation from research models to clinical decision support. These innovations collectively provide a foundation for scalable early detection and trial recruitment frameworks across the NHS and comparable health systems.

The use of routinely collected scans introduced challenges in cohort identification and image curation. These real-world clinical scans required exclusions for artefacts. All data access was within a TRE, where scans were pseudonymised and securely accessed. This setup required modifications to the ML development cycle^30^, including stringent disclosure controls^31^, ensuring data security and ethical use. This study focused on predicting future dementia diagnosis when brain changes are more likely to be subtle. In contrast, many high-accuracy studies^32^ predict dementia at a cross-sectional diagnostic stage, when structural abnormalities are pronounced. Furthermore, we used an age-matched dataset to ensure we were not just predicting older age. The field of dementia diagnosis using brain imaging has used various ML methods^33^, especially SVM, with some studies reporting higher accuracies^34–37^ than obtained in this study, but not when using comparable routinely acquired scan data, nor when predicting dementia diagnosis years in advance.

Our study further confirms that predictive sensitivity improves as the time to clinical diagnosis decreases, with peak sensitivity observed between 1.5 and 2 years prior. This aligns with earlier findings^38–40^ that show Alzheimer’s disease can be detected 3-4 years before clinical diagnosis. Our results support the idea of a “silent phase” in dementia progression, during which early structural brain changes (e.g., atrophy and white matter loss) occur before cognitive symptoms manifest^41^. Our accuracy levels are commensurate with a range of other studies reporting moderate to high dementia classification performance using a range of different risk models (not involving brain imaging) for predicting future dementia^32^.

The date of dementia diagnosis used in this study was derived from a validated algorithm^20^ which integrates information from multiple NHS data sources including primary care, secondary care, prescribing, and mortality records, to estimate when an NHS clinician first recognised dementia. Because no single gold-standard repository of dementia diagnoses exists within UK health data, this composite algorithm provides the best available real-world approximation of clinical diagnosis. Accordingly, predicting years in advance of this algorithm-derived date represents a genuine gain in lead time before dementia would be identified in real world NHS practice, creating a clinically meaningful window for early assessment or intervention.

Despite these promising findings, several limitations should be noted. As a feasibility study using a regional Scottish population, the sample size was limited, potentially affecting model generalizability. Routinely collected scans showed heterogeneity in image parameters and equipment, but this was necessary for scaling to a population level. The control group consisted of patients with non-dementia health conditions rather than a healthy population, potentially affecting specificity, but likely more realistic for clinical practice. We also grouped four dementia subtypes together for classification, which improved sample size but may have reduced subtype-specific insights. We did not account for time-to-diagnosis in model training, possibly masking temporal prediction dynamics. Future studies, such as the upcoming SCAN-DAN project^42^, will apply these cohort and data curation pipelines to a larger, nationwide Scottish dataset. This will help refine the model and validate its performance across more diverse populations.

There are currently no treatments that reverse Alzheimer’s or vascular dementia. The therapeutic goal is to slow or stop progression, making early prediction critical. Accurate identification of high-risk individuals enables timely intervention. For vascular dementia, existing strategies such as smoking cessation and blood pressure or lipid control can be implemented^43^. For Alzheimer’s disease, new disease-modifying drugs are emerging^44^, but their success depends on early, accurate patient identification. Given the high prevalence of dementia, prediction methods must use existing routinely acquired clinical scans, as collecting new data at scale is unfeasible at a population level. Many failed Alzheimer’s drug trials have cited the inability to identify patients early enough as a key reason for failure^45^. Our method addresses this inability and could help recruit large, well-characterised cohorts, for future trials of disease modifying drugs.

A key design choice in this feasibility study was to age- and sex-match dementia cases and controls, deliberately excluding age as a predictor despite its strong association with dementia risk. This approach isolated the independent predictive contribution of the brain scan itself, establishing proof of concept that routinely acquired MRI contains sufficient latent information to forecast future dementia. The intended next stage is to integrate this scan-derived probability and its distance-from-hyperplane confidence with readily available clinical variables, such as age, family history, and, where known, APOE genotype, to create a composite risk calculator analogous to those used in cardiology or oncology. In clinical use, a doctor could obtain a standard NHS brain scan, receive an automated binary prediction with confidence calibration, and then combine this with patient-specific information to derive an overall personalised dementia risk estimate for informed preventive care.

In summary, this study establishes that routinely acquired NHS brain MRI scans contain sufficient prognostic information to predict dementia years before it is clinically recognised. By using a validated, multi-source NHS algorithm to define diagnosis in the absence of a national gold standard, the model predicts meaningfully earlier than an NHS clinician would typically record dementia. The combination of routine imaging, real-world diagnostic timing, and confidence-calibrated prediction provides the foundation for a future composite risk calculator integrating scan-derived probability with readily available clinical variables. Such a system could transform dementia care from reactive diagnosis to proactive, population-scale prevention.

## Clinical and Research Implications

The results imply that predictive modelling of dementia risk can move beyond research datasets toward real-world, population-scale implementation. The ability to generate accurate individual patient predictions from routine NHS MRI scans further opens the opportunity of integrating machine-learning tools into existing radiology workflows and health-record systems, identifying high-risk individuals long before symptoms emerge. In practice, confidence-calibrated outputs such as the distance-from-hyperplane (DFH) measure could be embedded within reporting interfaces to flag cases where predictive certainty exceeds prespecified thresholds, supporting early preventive interventions or recruitment into disease-modifying trials before development of advanced disease.

From a health-system perspective, this approach provides a low-cost, scalable route to precision prevention, using data that are already being collected throughout the NHS. Future research should validate the method prospectively in larger, demographically diverse cohorts, assess temporal generalisability across sites, and integrate complementary risk factors such as genetics and blood biomarkers. If confirmed, the combination of routine NHS radiology, population linkage, and confidence-calibrated AI could enable a paradigm shift from symptomatic diagnosis to proactive, pre-symptomatic dementia prevention.

## Conclusion

In conclusion, this study presents a novel ML pipeline that utilizes routinely collected clinical scan data, showing considerable promise for predicting future dementia diagnosis, subtype prediction, and timeline forecasting. These findings lay the groundwork for future research, particularly the SCAN-DAN study, which will leverage the same cohort identification and data curation pipelines on a national scale. This expanded scope is expected to refine the model further, moving closer to a clinically translatable tool for predicting later dementia diagnosis with subtype differentiation.

## Supporting information

Supplementary Content

## Author contributions

All authors read and approved the final version of the manuscript. E.J., A.D. and D.S. conceived, designed and supervised the study. P.S.R and E.M implemented and validated the cohort identification pipeline, timeline window as conceived by A.D.. P.S.R implemented the image data curation pipeline, annotation tool and volume calculator under the guidance of D.S. and A.D.. The valid scans were manually curated, reviewed and validated by S.R. under the guidance of D.S. and A.D.. P.S.R. and E.M implemented the SPM12 based pre-processing under the guidance of D.S.. P.S.R implemented and validated the SVM based ML training testing conceived by D.S. This was further optimised by C.E.. S.A. performed formal analysis and investigation on classification results. P.S.R. and S.A. wrote the initial draft of the manuscript with assistance from S.S., S.K., A.D. and D.S.. The following drafts were reviewed and edited by P.S.R., S.A., C.E., S.R., E.M., S.S, M.R.K.M., S.K., E.T., E.J., A.D. and D.S. P.S.R. and S.A drafted the figures/graphs. P.S.R., S.K., E.J., A.D. and D.S. were involved in the planning and execution of this study. E.T., E.J., A.D. and D.S. acquired the financial support for this feasibility project leading to the work in this manuscript.

## Acknowledgements

Recruitment to this study was facilitated by SHARE - the Scottish Health Research Register and Biobank. SHARE is supported by NHS Research Scotland, Universities in Scotland and the Chief Scientists Office. The authors would like to acknowledge the eDRIS team (Public Health Scotland) and the Health Informatics Centre (HIC), University of Dundee for their support in obtaining approvals, data linkage, secure data access, and use of the Trusted Research Environment (TRE) which enabled this research. This work was supported by the Edinburgh International Data Facility (EIDF) and the Data-Driven Innovation Programme at the University of Edinburgh.

## Data sharing statement

The data related to the results presented in this article can be accessed within the HIC TRE subject to ethical and governance approvals. The details of dementia cohort identification can be accessed online on the HDR UK Phenotype library (https://phenotypes.healthdatagateway.org/phenotypes/PH1717/version/3973/detail/). The ML codebase used in this study is available online at GitHub (https://github.com/HicResearch/PICTURES-DementiaClassifier).

## Supplementary Content

*eMethod 1: Cases and controls identification using EMR datasets.*

*eMethod 2: Identifying valid brain scans*

*eMethod 3: Preprocessing valid brain scans*

*eMethod 4: Feature reduction*

*eMethod 5: Hyper-parameter details*

*eMethod 6: Patient cohort identified using EMR datasets*

*eTable 1: Selection of ICD-9 and ICD-10 codes used to select different dementia types.*

*eFigure 1: Example of time window showing the timeline of events used for selecting valid scans.*

*eFigure 2: Snapshot showing GUI of bespoke annotation tool developed in RShiny and used to manually inspect and annotate brain scans.*

*eFigure 3: Flowchart showing the volume calculation to filter partial brain scans*

*eFigure 4: Summary of sequence parameters for T1w MRI images. Where TR: repetition time, TE: echo time.*

*eFigure 5: Number of voxels selected during feature reduction in 5-fold nested cross-validation*

*eFigure 6: Heatmap showing grid search performance across hyperparameters.*

