## Supplementary Content for "Machine learning-based prediction of future dementia using routine clinical MRI brain scans and healthcare data"

### ***eMethod 1:*** Cases and controls identification using EMR datasets

**Dementia Cases**

Cases were selected from GoDARTS and SHARE studies using the following four linked datasets:

1. **Dispensed prescribing**: This dataset includes the detailed longitudinal prescription records of all medications that are dispensed from the pharmacies to the individuals. The drugs specific to dementia i.e. Chapter 4.11 of the British National Formulary (BNF) were identified.
2. **Death**: The Scottish General Register Office (GRO) records information of death certification. The ICD-9 and ICD-10 codes related to dementia were identified in these certifications (See Appendix Table 1).
3. **General hospitalisation admissions (SMR01)**: The general hospital inpatient and day case discharge information include the individual episode level data. The ICD-9 and ICD-10 codes related to dementia were identified in these records (See Appendix Table 1).
4. **Psychiatric hospital discharge (SMR04)**: The psychiatric hospital inpatient and day case discharge information include the individual episode level data. The ICD-9 and ICD-10 related to dementia were identified from these records (See Appendix Table 1).

For each patient with dementia, the incident date of dementia was evaluated based on the earliest date of occurrence in any of these datasets. As EMR records are independently coded episodes over time, patients could receive codes from different dementia subtypes due to the intrinsic inaccuracy in the clinical coding. A probabilistic model was used[1] to assign a dementia subtype to such patients.

**Non-Dementia Controls**

These patients were selected from GoDARTS and SHARE studies and have:

1. No dementia specific drugs (Chapter 4.11 of the BNF) in the dispensed prescribing records, and
2. No ICD-9 and ICD-10 codes related to dementia in death, general hospital admissions, and psychiatric hospital discharge datasets.

The algorithm for identifying the dementia cases and non-dementia controls was implemented in R[2] using DBI[3], dplyr[4], string[5] and odbc[6] packages.

### ***eMethod 2:*** Identifying valid brain scans

A valid scan for the purpose of this study was defined as a T1 weighted volume of the entire brain, without significant artefact or pathological structural distortion (e.g. large stroke or large tumour). Firstly, the DICOM metadata was searched for text strings ‘T1’ AND ‘MR’ AND ‘Head’ sequence type scans. A time window approach is then used to select the valid scans. Valid case scans were identified as scans acquired one to six years before the earliest date of diagnosis of dementia in their medical record. Similarly, valid control scans were identified as scans acquired 6 years before the last date of follow-up or date of death. The valid images are identified using the following time window (See Appendix Figure 1):

*For valid dementia patient scans:*

*(edd - ds) ≥ 1yr AND (edd - ds) ≤ 6 yrs*

*For valid control patient scans:*

*(ldf – ds) ≥ 6 yrs*

*Where, ds* is date of scan, *edd* is the earliest date of diagnosis for dementia patient *and ldf* is the latest date of follow-up for non-dementia control patients.

Next, a bespoke ‘annotation tool’ was developed and used to manually inspect the anatomical view in all three planes (axial, sagittal, and coronal) of these remaining scans along with their DICOM metadata. This manual review was useful to exclude localizer survey scans (with <10 2D slices), partial brains (identified using bespoke volume calculation, See Appendix Note 3 and Appendix Figure 4), brains with large lesions/tumours and other incorrectly tagged body part scans.

In order to filter partial brain scans, each valid scan (having number of slices > 10) is read from the memory to capture the orthographic view (See Appendix Figure 3). Finally, the value of volume V is calculated (in $cm^3$) as follows:

$$\mathbf{V}(MRAquisitionType_{(0018,0023)}) = \begin{cases} (SpacingBetweenSlices_{(0018,0088)} * noofSOPInstances * noofRows * noofColumns * PixelSpacing^2_{(0028,0030)})/1000 & MRAquisitionType_{(0018,0023)} = 2D \\ (SliceThickness_{(0018,0050)} * noofSOPInstances * noofRows * noofColumns * PixelSpacing^2_{(0028,0030)})/1000 & MRAquisitionType_{(0018,0023)} = 3D \end{cases}$$

The orthographic view and the corresponding volume is displayed in the RShiny-based annotation tool.

The final list of valid scans was then saved. To reduce gender and age confounds potentially arising through the IDCP process, the number of males and females as well as their age were balanced between the control and the patient groups, such that both groups had equal proportions of aged-matched males and females. Subsequently, the ‘annotation tool’ was used to manually review the matched control scans for exclusion of partial brain views, large lesions/tumours and other anatomical abnormalities. In case of any partial brains/lesions present in control scan, an intermediate list was saved, and the age sex matching performed again to obtain another valid control scan from the pool. Finally, the updated matched cohort of ‘normal’ (without abnormalities) whole brain scans were converted from dicom to nifti format for SPM[7] software usage. The IDCP, annotation tool and volume calculation were implemented in R Language[2] using DT[8], plyr[9], oro.dicom[10], oro.nifti[11], neurobase[12], future.apply[13] and shiny[14] packages.

### *eMethod 3:* Preprocessing valid brain scans

All T1-weighted magnetic resonance scans from IDCP were pre-processed using SPM12[7], running under MATLAB™ R2021a[15]. MRI scans were bias-field corrected, spatially normalised to the MNI template and segmented creating spatially normalized images of different brain 'tissue' types: grey matter (GM), white matter (WM), and cerebrospinal fluid (CSF).

Following tissue segmentation, a nonlinear within-subject registration of the segmented GM and WM tissues was performed using the SPM12 Geodesic Shooting Toolbox[16]. This involves iteratively refining a template created from the averaged of all subjects' segmented GM and WM images, computing deformations between the template and segmented subjects' images, applying inverse deformations, and averaging the results to update the template. This continues until the template accurately represents common features across all the input images.

The *scalar momentum*[17] features were extracted that encodes the deformation fields associated with the segmented GM and WM tissues, and also the residual between these tissue types and the template, as defined by equation 1-3 below:

$|D\phi|(\mu 1 - c1(\phi))$................................(1)

$|D\phi|(\mu 2 - c2(\phi))$................................(2)

$|D\phi|(c1(\phi) + c2(\phi) - \mu 1 - \mu 2)$.................(3)

Where:

- $\phi$ represents the forward transformation field
- $|D\phi|$ denotes the Jacobian determinants of $\phi$
- $\mu 1$ is the GM template
- c1 represents GM
- $\mu 2$ is the WM template
- c2 represents WM

This captures anatomical variability, tissue density differences, morphological changes and local geometric features, providing a comprehensive representation of the brain's structure,[18] which has found to be empirically useful for maximising ML performance[18].

### *eMethod 4:* Feature reduction

Feature reduction to avoid over fitting due to the ‘curse of dimensionality’, it is important to select a subset of informative voxels and to discard noisy (random variation) and redundant voxels[19]. Successful feature reduction improves the performance of the classification and reduces computational cost[19,20]. The present study employed a voxel-wise two sample t-test embedded within a nested cross-validation SVM framework[21]. This framework is very flexible and not specific to particular illnesses, having been used for depression[22], non-brain structure fMRI data[23], and non-neuroimaging cognitive data[24].

The t-test is computationally fast, easy to implement and scales well to high dimensional data[19]. To avoid double-dipping and overfitting the model, the univariate t-map was computed using only the training set data[25]. The t-map represents the t-statistic value for each voxel, which indicates the statistical significance of the difference between the two groups. The t-map was then used to create a t-mask, which is a binary mask that selects only the voxels with a p-value below an automatically selected threshold. This p-value threshold indicates that these voxels are statistically significant for training purposes. this threshold being optimized at the same time as the classifier's hyperparameters using nested cross-validation. The t-masks were applied to the testing set to evaluate the classification performance at different p-value thresholds to improve model generalizability.

### *eMethod 5:* Hyper-parameter details

In this study, a radial basis function (RBF)-SVM, as implemented in MATLAB, was employed[26]. SVM is one of the most widely used supervised ML methods in Multi-Variate Pattern Analysis (MVPA) for neuroimaging studies, due to its effectiveness in dealing with high-dimensional datasets and its flexibility in decoding diverse sources of brain data[27]. RBF-SVM offer better performance on small datasets in comparison to deep learning models such as Convolutional Neural Networks[28].

A nested cross-validation approach was applied to optimise two parameters: the p-value threshold of the t-test feature reduction, and the box-constraint “C”, which determines the extent to which wrong predictions are penalised. The predefined range of values for each hyperparameter was as follows: p-value [0.000001, 0.00001], kernel width ‘λ’ [0.001, 200], and C [0.001, 200], were tested using evenly spaced values within these ranges. These parameters were tested using SLURM on cloud-based TRE[29].

Our feature reduction t-test was applied to the training subset of the inner folds, generated multiple binary t-masks for each p-value threshold, to filter voxels for inclusion in the subsequent ML training and testing.

Feature reduction and RBF-SVM used a combination of hyperparameters, with model performance evaluated on the left-out inner fold. This procedure was repeated such that each inner fold served as the validation set once, and the combination of hyperparameters that yielding the highest average classification accuracy across the inner folds was selected. This optimal set of hyperparameters was then used for training and testing of the RBF-SVM on the outer fold. This entire procedure was then repeated for each outer fold, ensuring that each fold was used as a test set once. The overall accuracy was calculated by averaging the classification accuracies across all outer folds.

### ***eMethod 6:*** Patient cohort identified using EMR datasets

**Dementia Patients**

Following count of patients were selected from GoDARTS and SHARE studies using the following four linked datasets:

1. **Dispensed prescribing**: Patients identified [n=1932].
2. **Death**: Patients identified: Alzheimer's Dementia [n=513], Vascular Dementia [n=572], Unspecified Dementia [n=435], and Other Dementia [n=42]. Total records [1562] and total common patients [1463].
3. **General hospitalisation admissions (SMR01)**: Patients identified: Alzheimer's Dementia [n=791], Vascular Dementia [n=759], Unspecified Dementia [n=869], and Other Dementia [n=353]. Total records [n=2772] and total common patients [n=2219].
4. **Psychiatric hospital discharge (SMR04)**: Patients identified: Alzheimer's Dementia [n=208], Vascular Dementia [n=131], Unspecified Dementia [n=101], and Other Dementia [n=30]. Total records [n=470] and total common patients [n=411].

Overall, 3477 dementia patients were identified which included Alzheimer's Dementia [n=1858], Vascular Dementia [n=800], Unspecified Dementia [n=617], and Other Dementia [n=202].

**Non-Dementia Control Patients**

In total, 99105 non-dementia patients were identified from the GoDARTS and SHARE studies which neither have any dementia related ICD-9 and ICD-10 codes nor prescription BNF codes.

***eTable 1:*** Selection of ICD-9 and ICD-10 codes used to select different dementia types

| **Alzheimer's Disease (AD)** | **Vascular Dementia (VD)** | **Unspecified Dementia (UD)** | **Other Dementia (OD)** |
|---|---|---|---|
| ^3310 | ^2904 | ^2900 | ^2912 |
| F00 | F01 | ^2901 | ^2982 |
| G30 | | ^2902 | ^2941 |
| | | ^2903 | ^3311 |
| | | ^2942 | ^3312 |
| | | ^3312 | ^33119 |
| | | F03 | ^33111 |
| | | G311 | ^33182 |
| | | | ^29282 |
| | | | F02 |
| | | | A810 |
| | | | F051 |
| | | | G310 |
| | | | G318 |
| | | | ^0461 |
| | | | ^797 |
| | | | F1027 |
| | | | F1097 |
| | | | G3110 |
| | | | G3109 |
| | | | G3183 |

***eFigure 1:*** Example of time window showing the timeline of events used for selecting valid scans.

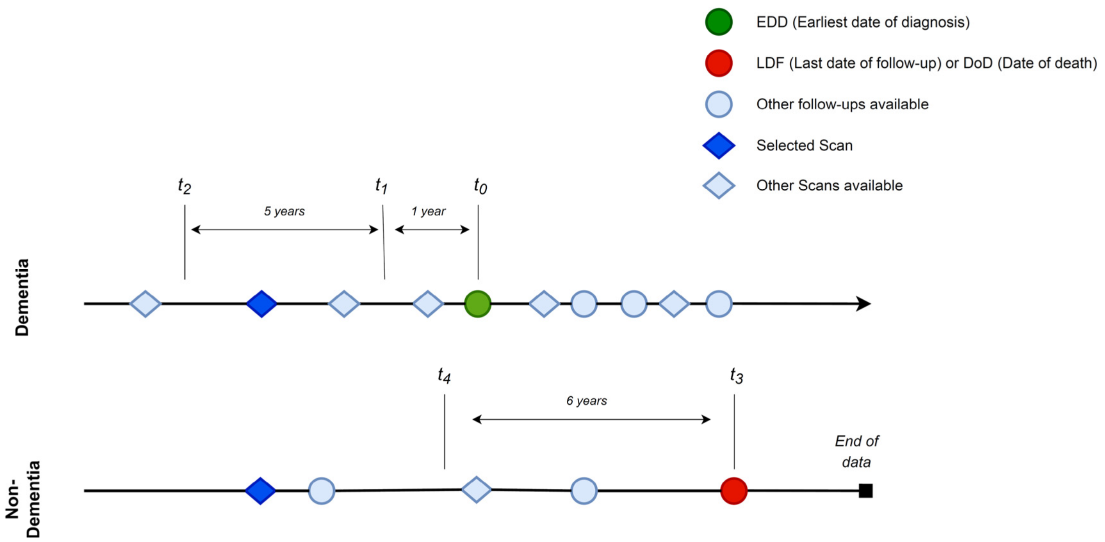

***eFigure 2:*** Snapshot showing GUI of bespoke annotation tool developed in RShiny and used to manually inspect and annotate brain scans.

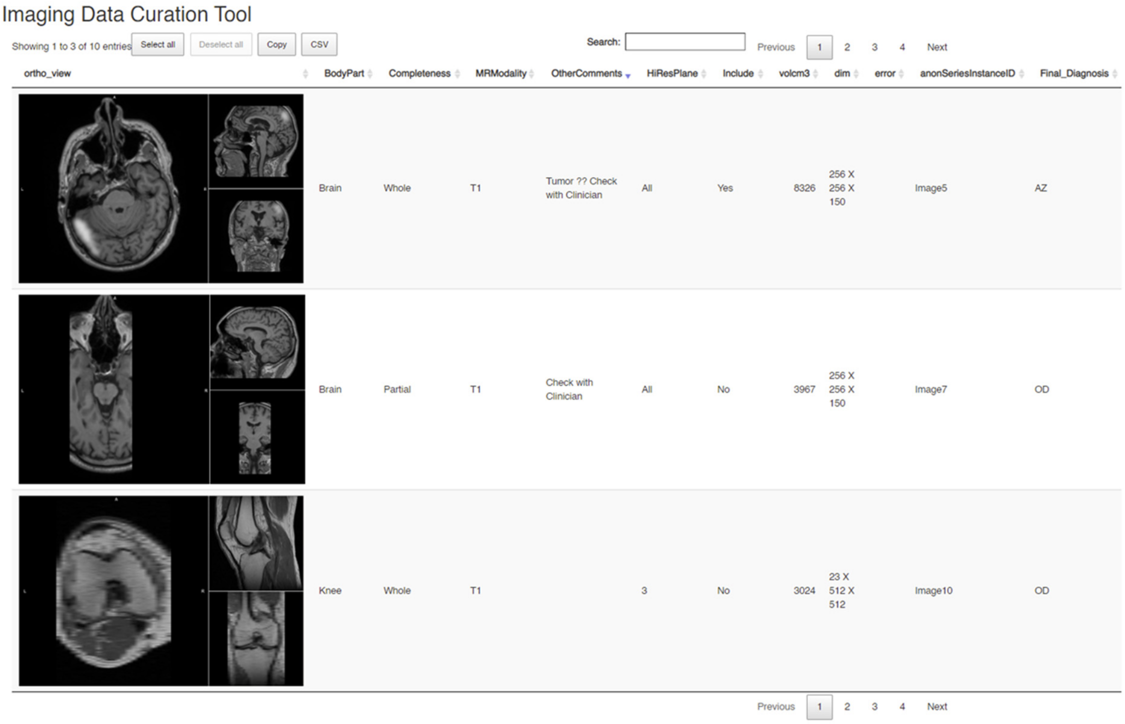

***eFigure 3:*** Flowchart showing the volume calculation to filter partial brain scans.

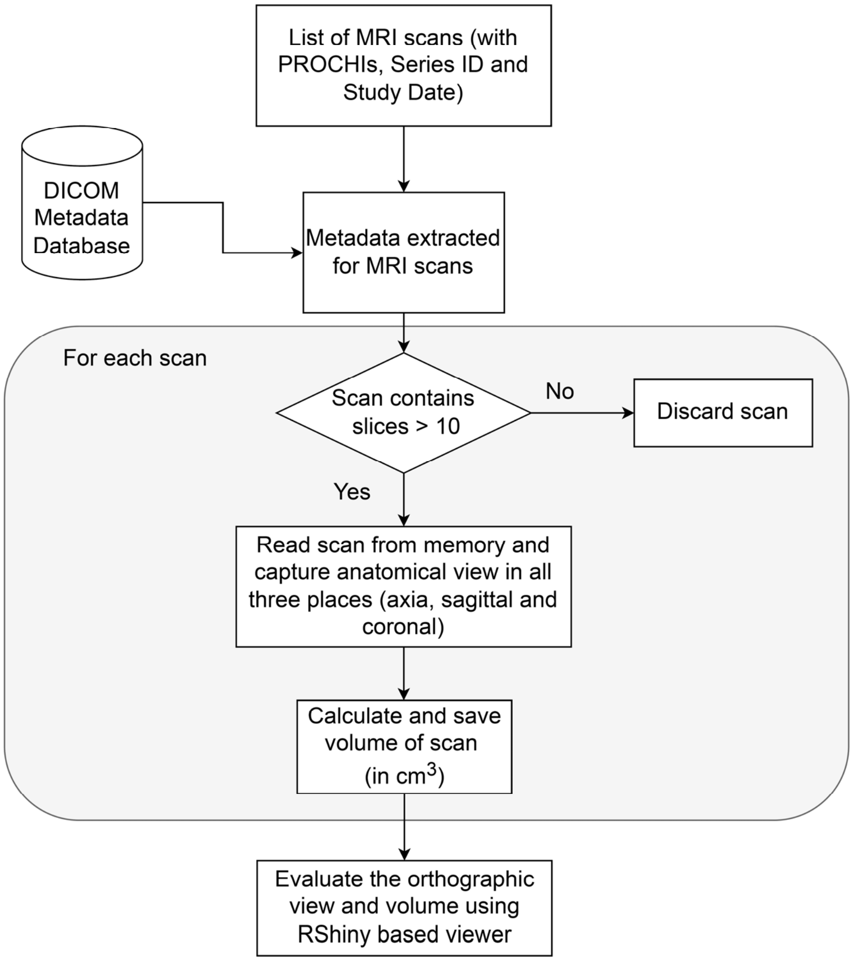

***eFigure 4:*** Summary of sequence parameters for T1w MRI images. Where TR: repetition time, TE: echo time.

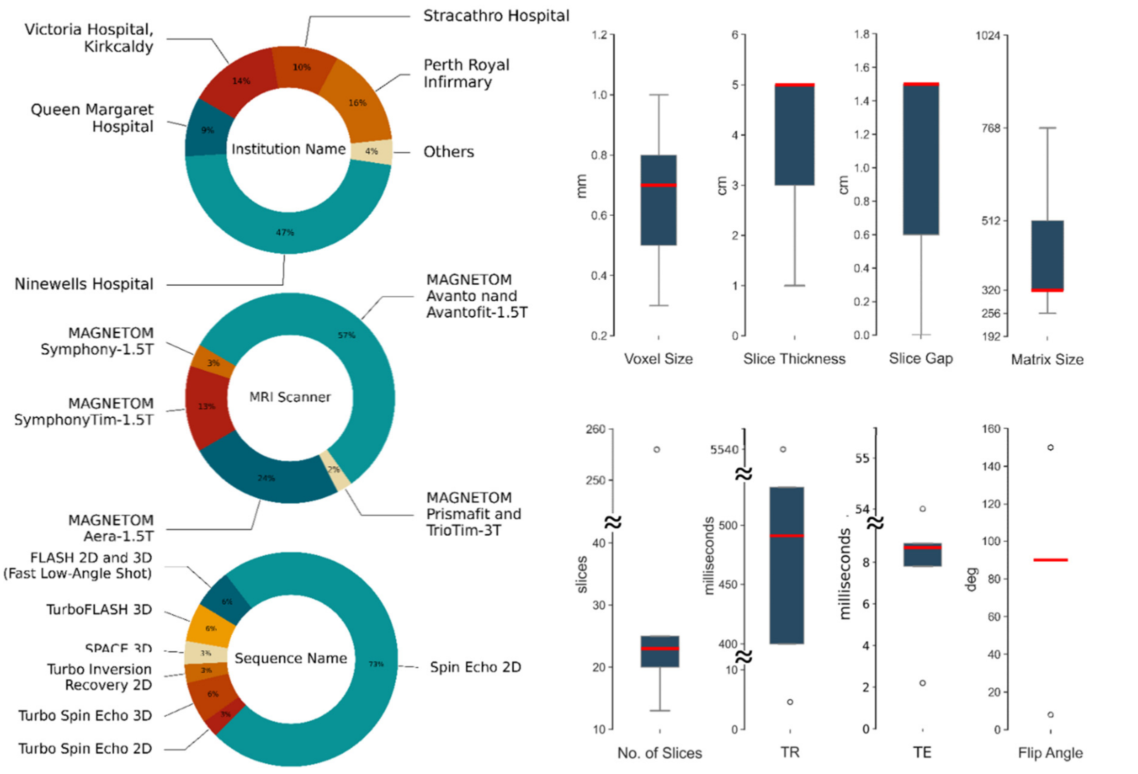

***eFigure 5:*** *Number of voxels selected during feature reduction in 5-fold nested cross-validation*

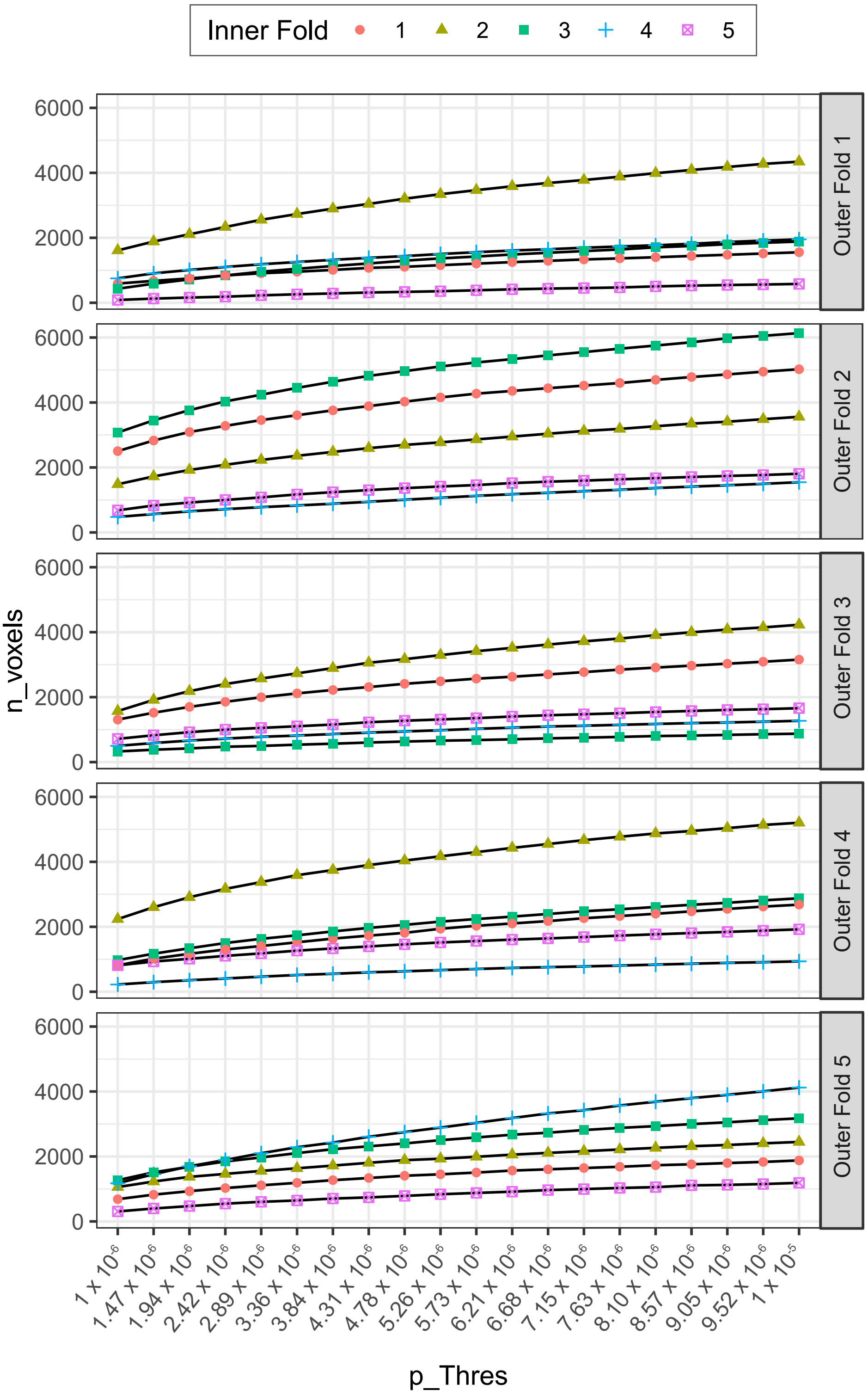

*eFigure 6: Heatmap showing grid search performance across hyperparameters*

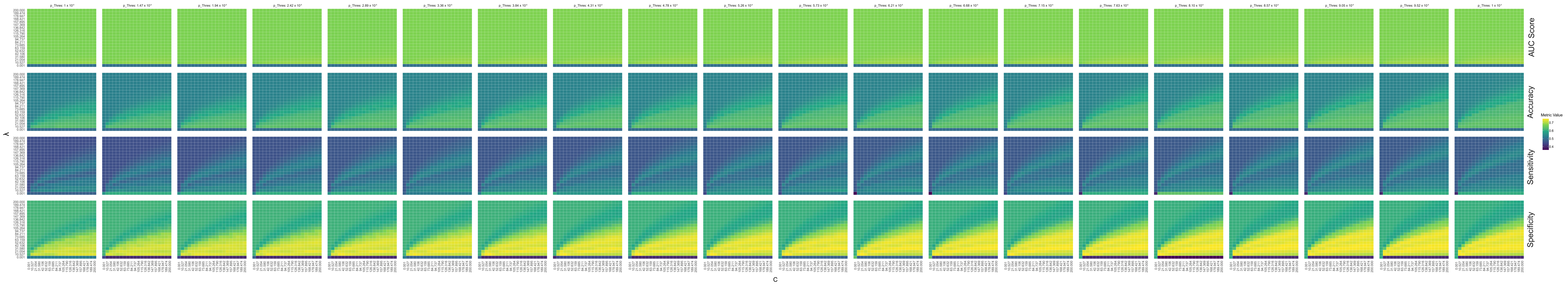
